# Trajectories of multiple long-term conditions and mortality in older adults: A retrospective cohort study using English Longitudinal Study of Ageing (ELSA)

**DOI:** 10.1101/2023.05.18.23290151

**Authors:** Christos V. Chalitsios, Cornelia Santoso, Yvonne Nartey, Nusrat Khan, Glenn Simpson, Nazrul Islam, Beth Stuart, Andrew Farmer, Hajira Dambha-Miller

**Affiliations:** Primary Care Research Centre, University of Southampton, Southampton, UK; Centre for Evaluation and Methods, Wolfson Institute of Population Health, Queen Mary University of London, London, UK; Nuffield Department of Primary Care Health Sciences, University of Oxford, UK

**Keywords:** Multiple long-term conditions, trajectories, mortality, English Longitudinal Study on Ageing (ELSA), older adults

## Abstract

**Objectives:** To classify older adults with MLTC into clusters based on accumulating conditions as trajectories over time, characterise clusters and quantify associations between derived clusters and all-cause mortality.

**Design:** We conducted a retrospective cohort study using the English Longitudinal Study of Ageing (ELSA) over nine years (n=15,091 aged 50 years and older). Group-based trajectory modelling was used to classify people into MLTC clusters based on accumulating conditions over time. Derived clusters were used to quantify the associations between MLTC trajectory memberships, sociodemographic characteristics, and all-cause mortality.

**Results:** Five distinct clusters of MLTC trajectories were identified and characterised as: “no-LTC” (18.57%), “single-LTC” (31.21%), “evolving MLTC” (25.82%), “moderate MLTC” (17.12%), and “high MLTC” (7.27%). Increasing age was consistently associated with an increased number of MLTC. Female sex (aOR = 1.13; 95%CI 1.01 to 1.27) and ethnic minority (aOR = 2.04; 95%CI 1.40 to 3.00) were associated with the “moderate MLTC” and “high MLTC” clusters, respectively. Higher education and paid employment were associated with a lower likelihood of progression over time towards an increased number of MLTC. All the clusters had higher all-cause mortality than the “no-LTC” cluster.

**Conclusions:** The development of MLTC and the increase in the number of conditions over time follow distinct trajectories. These are determined by non-modifiable (age, sex, ethnicity) and modifiable factors (education and employment). Stratifying risk through clustering will enable practitioners to identify older adults with a higher likelihood of worsening MLTC over time to tailor effective interventions.

**Strengths and limitations:**

- The main strength of the current study is the use of a large dataset, assessing longitudinal data to examine MLTC trajectories and a dataset that is nationally representative of people aged 50 years and older, including a wide range of long-term conditions and sociodemographics.
- The measurement of MLTC was limited to ten long-term conditions, which was all of what was available in the English of Longitudinal Study of Ageing, which may not be exhaustive of all possible long-term conditions.

## Introduction

Globally, the average life expectancy has risen from 66.8 years in 2000 to 73.4 years in 2019 (1). By 2050, the population over 60 and 80 years will reach 2.1 billion and 426 million, respectively (2,3). This rise in longevity raises the risk of developing multiple long-term conditions (MLTC), which is the co-occurrence of two or more chronic diseases (4). Globally, the prevalence of MLTC among older people is reported to be between 55-98% (5), and in the UK, this is expected to rise from 54% in 2015 to 68% in 2035 (2). MLTC represent an ongoing challenge for healthcare systems because people with MLTC have worse care outcomes, including functional limitation and disability (6,7), higher service utilisation (5), mortality (8) and poorer quality of life (5). Management of MLTC places considerable economic and logistical burdens on services which are traditionally organised around single disease models (6).

While there is ample evidence of identified risk factors (7,9) and adverse care outcomes for MLTC cross-sectionally to help understand the prevalence and patterns of MLTC, they provide little evidence on temporal elements, including patterns of MLTC development over time (8,10,11). There is a paucity of longitudinal approaches examining patterns in the accumulation of diseases over time (12). Understanding the trajectory that an older adult will follow in the progression towards an increased number of MLTC could help predict when intervention is needed and inform targeted and earlier preventive interventions. To address this critical gap in the literature, this study aimed to classify older adults with MLTC into clusters based on the cumulation of conditions as trajectories over time; clusters were then characterised, and associations were quantified between derived clusters and all-cause mortality.

## Methods

### Data source and study population

The English Longitudinal Study of Aging (ELSA) is a longitudinal cohort of people aged 50 years or older living in England (13). The ELSA cohort profile has been described in detail elsewhere (14). In summary, it included 12,099 people at study entry in 2002 with follow-up every two years with self-report questionnaires on physical and mental health, well-being, finances, and attitudes around ageing over time. Four yearly additional nurse visits collected objective data such as anthropometric data (13,15). The ELSA is an open cohort, and refreshment samples were added depending on the proportional age requirement for ELSA, so the total number of people in this cohort was 15091. Our baseline was wave 2 (2004/5) of the ELSA cohort, the first collecting time point in the study of long-term conditions with a nine-year follow-up to wave 6 (2012/3), the most recent wave with available data on all-cause mortality status.

### Multiple Long-Term Conditions

MLTC was defined as the presence of two or more of the following ten conditions: hypertension, diabetes, cancer, lung disease, cardiovascular disease, stroke, mental health disorder, arthritis, Parkinson’s disease, and dementia. These are self-reported by patients, relatives or carers and verified by nurse visits (13). These ten conditions were available within the ELSA dataset based on our earlier work to define MLTC (16,17). After statistical consideration due to the small sample size and clinical discussion, we grouped some of the conditions as follows: people with depression were combined with mental health disorders, asthma was combined with lung disease, Alzheimer’s within dementia, and finally, those with heart attack, angina, heart murmur, abnormal heart rhythm and congestive heart failure combined into those with cardiovascular disease.

### All-cause mortality

All-cause mortality was reported by end-of-life interviews on waves 2, 3, 4 and 6 with relatives and friends after death.

### Covariates

Sociodemographic variables included were age, sex, ethnicity (defined as white/non-white), education, employment, and marital status. The education variable was categorised into four groups: less than upper secondary level, upper secondary and vocational level, tertiary level, and others. Employment status was categorised into ‘paid employment and ‘unemployed’. Marital status was categorised into three groups: never married, married/having a partner, and separated/divorced/widowed. These covariates were based on the baseline. We used data provided in the nearest subsequent waves if they were missing at baseline.

### Statistical analysis

Descriptive statistics were used to summarise participants’ characteristics. We used group-based trajectory modelling (GBTM) to classify older adults with MLTC into clusters based on the accumulation of conditions as trajectories over time. GBTM is a finite mixture model applying maximum likelihood to identify a cluster of people following similar trajectories by the number of conditions over time (18). This model assumes the same error variance for all clusters and time points and treats missing data as ‘missing at random’ (19). The procedure for selecting the best model included two steps: identifying the ideal number of trajectory groups and determining polynomial orders to represent the shapes of the trajectories (18,20). Based on the observed distribution, we employed a censored normal model to specify MLTC (21,22). We fitted the models iteratively, starting with one and increasing up to a maximum of six clusters that would be useful in a clinical setting (20). We selected the number of trajectory clusters based on the following criteria: the lowest Bayesian Information Criterion (BIC) value, Average Posterior Probability Assignment (APPA) >70%, Odds of a Correct Classification (OCC) >5, the percentage of participants in each trajectory groups >5% of the total sample (if less than 5% it is unlikely to be conceptually useful for clinical practice) (22–24). We first used cubic polynomials to characterise the shape of the clusters of MLTC trajectories. However, after selecting the number of trajectories, we refitted the model to use lower-order terms when the higher-order terms were insignificant (20). We then assigned individuals to the trajectory group based on the maximum posterior probability (20). Multinomial logistic regression was then performed to test the association between socio-demographic factors and clusters of MLTC trajectory, with the “no-LTC” cluster as the reference. Binary logistic regression was also performed to quantify the association between the clusters of MLTC trajectory membership and all-cause mortality, adjusting for all the covariates mentioned above. A squared term of age was included in the model to account for the non-linear relationship between age and mortality. The significance level was set at a p-value <0.05, and all analyses were performed using STATA M.P v17.0.

### Patient and Public Involvement

This study was conducted as part of a wider mixed-methods programme of research exploring the potential of machine learning to address multimorbidity through the ‘clustering’ of patients based on similarities in clinical and social care needs. Patient and public involvement has been incorporated throughout the wider research programme from the initial inception, design, and dissemination of findings. The initial results and the final written draft of the study submitted in this manuscript were shared with our programme’s patient and public representative.

## Results

### Participants’ characteristics

We identified 15,091 individuals participating in at least one wave during the follow-up period (The flow of participants through the study is shown in **Figure 1**). Six participants were excluded, as they had no information on MLTC. After excluding those (n = 123) with missing data on covariates, 14,962 people were included in the final analysis. The mean (SD) age of the cohort was 61.9 (11) years; most were females (53.5%), whites (96.5%), with educational attainment of upper secondary or vocational (43.1%), employed (56.8%), and married or had a partner (72%) (**Table 1**).

**Figure 1.**
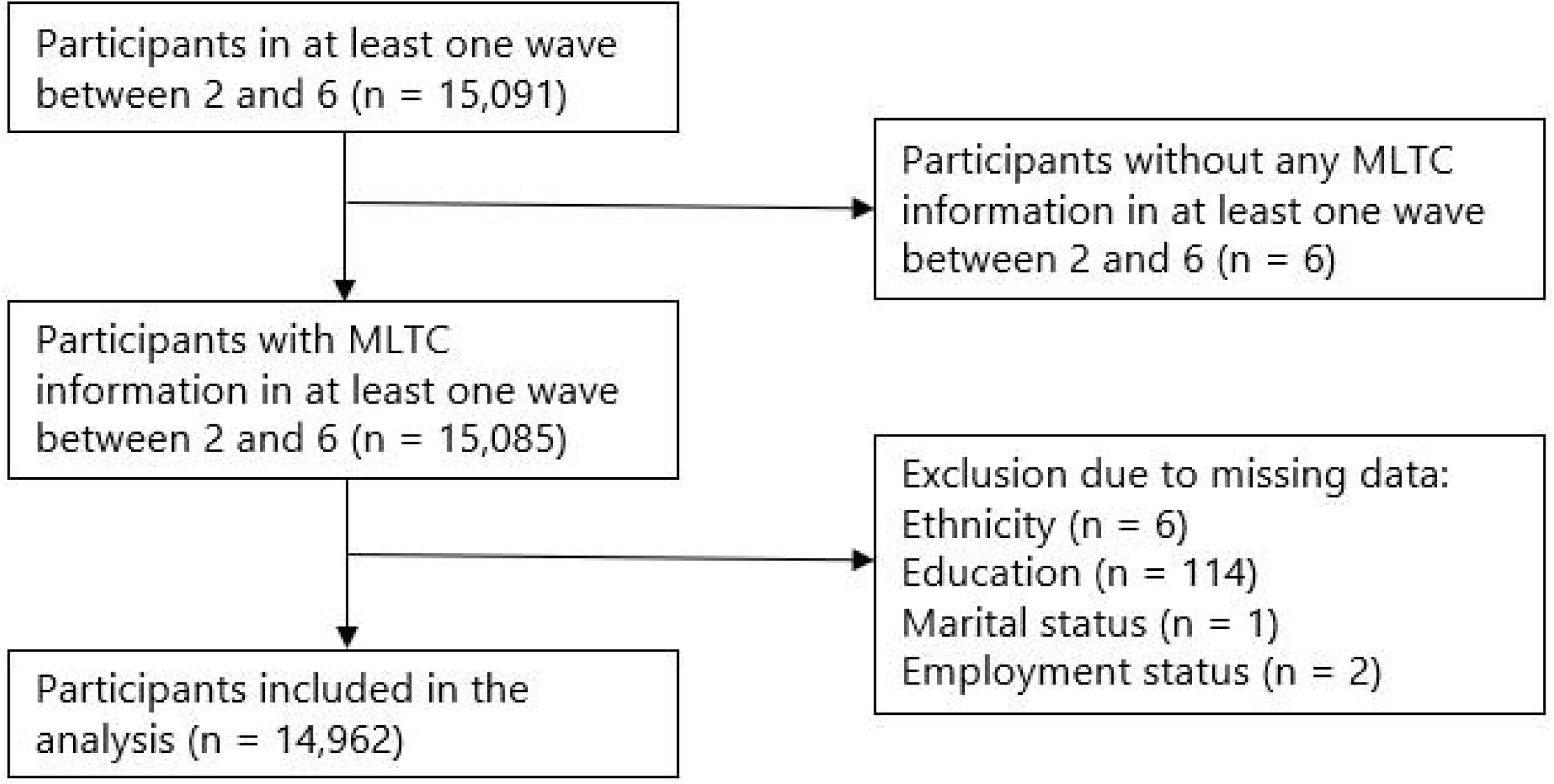
Flow chart of participants selection. MLTC, multiple long-term conditions.

**Table 1.** Participants’ characteristics overall and stratified by clusters of MLTC trajectory.

|  | **Total** | **No-LTC** | **Single-LTC** | **Evolving MLTC** | **Moderate MLTC** | **High MLTC** |
| --- | --- | --- | --- | --- | --- | --- |
|  | 14962 (100%) | 2826 (18.9%) | 4802 (32.1%) | 3739 (25.0%) | 2532 (16.9%) | 1063 (7.1%) |
| **Age**, mean (SD) | 61.9 (11) | 56.0 (9.1) | 60.0 (10.0) | 62.9 (10.8) | 67.1 (10.7) | 69.8 (10.4) |
| **Sex** |  |  |  |  |  |  |
| Male | 6951 (46.5) | 1402 (20.2) | 2361 (34.0) | 1675 (24.1) | 1050 (15.1) | 463 (6.7) |
| Female | 8011 (53.5) | 1424 (17.8) | 2441 (30.5) | 2064 (25.8) | 1482 (18.5) | 600 (7.5) |
| **Ethnicity** |  |  |  |  |  |  |
| White | 14440 (96.5) | 2726 (18.9) | 4629 (32.1) | 3618 (25.1) | 2451 (17.0) | 1016 (7.0) |
| Non-white | 522 (3.5) | 100 (19.2) | 173 (33.1) | 121 (23.2) | 81 (15.5) | 47 (9.0) |
| **Education** |  |  |  |  |  |  |
| Less than upper secondary | 5107 (34.1) | 629 (12.3) | 1417 (27.8) | 1326 (26.0) | 1136 (22.2) | 599 (11.7) |
| Upper secondary, vocational | 6444 (43.1) | 1399 (21.7) | 2186 (33.9) | 1609 (25.0) | 941 (14.6) | 309 (4.8) |
| Tertiary | 2277 (15.2) | 626 (27.5) | 859 (37.7) | 497 (21.8) | 227 (10.0) | 68 (3.0) |
| Others | 1134 (7.6) | 172 (15.2) | 340 (30.0) | 307 (27.1) | 228 (20.1) | 87 (7.7) |
| **Employment** |  |  |  |  |  |  |
| Paid employment | 8500 (56.8) | 895 (10.5) | 2278 (26.8) | 2333 (27.5) | 2033 (23.9) | 961 (11.3) |
| Unemployed | 6462 (43.2) | 1931 (30.0) | 2524 (39.1) | 1406 (21.8) | 499 (7.7) | 102 (1.6) |
| **Marital status** |  |  |  |  |  |  |
| Never married | 789 (5.3) | 148 (18.8) | 268 (34.0) | 189 (24.0) | 131 (16.6) | 53 (6.7) |
| Married/partner | 10766 (72.0) | 2282 (21.2) | 3635 (33.8) | 2674 (24.8) | 1566 (14.6) | 609 (5.7) |
| Separated/divorced/widowed | 3407 (22.8) | 396 (11.6) | 899 (26.4) | 876 (25.7) | 835 (24.5) | 401 (11.8) |

### Clusters of MLTC trajectory

We examined one to six clusters in the model to determine the optimal cluster number. Five clusters were selected based on the model fit indicators (**Supplementary Table 1**) and the interpretability of classified trajectories.

Participants displayed high posterior probabilities of belonging to their assigned clusters ranging from 0.88 to 0.97 across the five clusters. The “no-LTC” cluster (18.57%) was dominated by people (95.2%) without any record of the examined long-term condition during the follow-up, and the “single-LTC” cluster (31.21%) consisted of those who did not develop MLTC during the study period but may have had one long-term condition (**Figure 2**). The “evolving MLTC” cluster (25.82%) was characterised by people who progressed from less than two long-term conditions at baseline to two, three, or four by the end of follow-up. Two clusters had MLTC profiles which showed increasing numbers of long-term conditions (“moderate MLTC” (17.12%) and “high MLTC” (7.27%)). Those in these clusters started with MLTC and continued to have higher counts of long-term conditions in the following periods.

**Figure 2.**
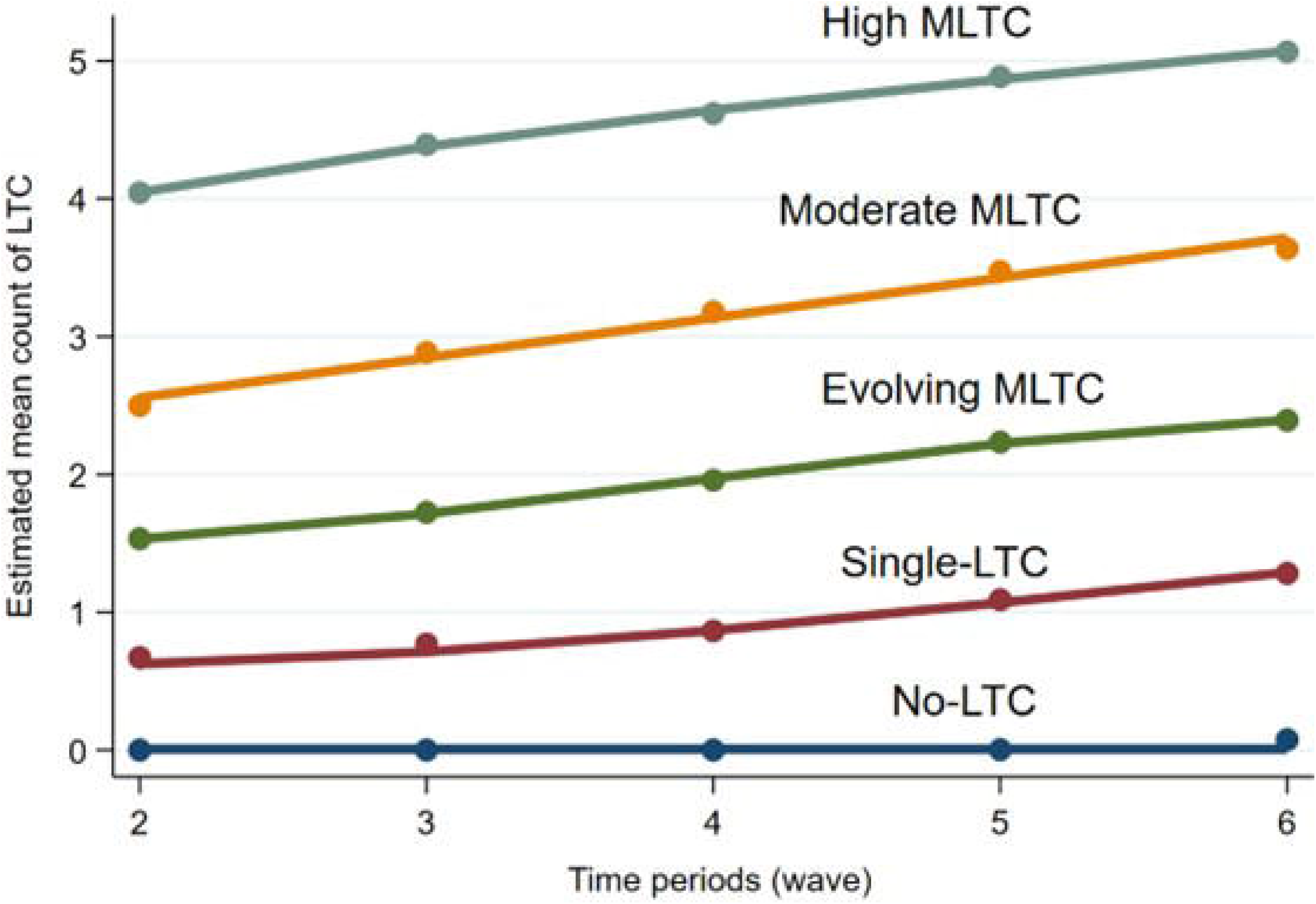
Clusters of MLTC trajectories over time (wave 2 to 6) in the English Longitudinal Study of Aging (ELSA) study. The solid lines represent the estimated mean count of MLTC profiles for the five clusters. The “no-LTC” cluster included people who did not have any of the examined long-term conditions; the “single-LTC” cluster included those who did not develop MLTC but may have had one long-term condition; the ‘‘evolving MLTC’’ cluster included those who developed MLTC lately; the ‘‘moderate MLTC’’ cluster included those who started with the lower number of MLTC and developed further long-term conditions; the ‘‘high MLTC’’ cluster consisted of those who started with the higher number of MLTC and developed additional long-term conditions. Abbreviation: MLTC, Multiple long-term conditions.

### Clusters of MLTC trajectory and socio-demographic characteristics

Increasing age was consistently associated with all MLTC clusters, compared to the “no-LTC” cluster (**Table 1 & 2**). Females had higher odds (aOR = 1.13; 95%CI 1.01 to 1.27) of being in the “moderate MLTC” clusters than females. Being non-white increased the odds of belonging to the “high MLTC” cluster by 2.04 times (aOR = 2.04; 95%CI 1.40 to 3) compared to whites. Higher education and paid employment decreased the odds of belonging to any of the four clusters than those with less than upper secondary education and unemployment, respectively.

**Table 2.** The association between socio-demographic factors and clusters of MLTC trajectories.

| Socio-demographics | Adjusted OR (95%CI) (Reference: No-LTC) |  |  |  |
| --- | --- | --- | --- | --- |
|  | Single-LTC | Evolving MLTC | Moderate MLTC | High MLTC |
| **Age** | 1.04 (1.03-1.04) | 1.05 (1.05-1.06) | 1.07 (1.06-1.08) | 1.08 (1.07-1.09) |
| **Sex** |  |  |  |  |
| Male | Reference | Reference | Reference | Reference |
| Female | 1.00 (0.91-1.10) | 1.11 (0.99-1.23) | 1.13 (1.01-1.27) | 0.95 (0.81-1.11) |
| **Ethnicity** |  |  |  |  |
| White | Reference | Reference | Reference | Reference |
| Non-white | 1.17 (0.91-1.50) | 1.13 (0.85-1.49) | 1.36 (1.00-1.86) | 2.04 (1.40-3.00) |
| **Education** |  |  |  |  |
| Less than upper secondary | Reference | Reference | Reference | Reference |
| Upper secondary, vocational | 0.92 (0.81-1.03) | 0.87 (0.77-0.99) | 0.77 (0.67-0.88) | 0.53 (0.45-0.64) |
| Tertiary | 0.84 (0.72-0.97) | 0.68 (0.58-0.80) | 0.51 (0.42-0.62) | 0.33 (0.25-0.45) |
| Others | 1.01 (0.83-1.25) | 1.04 (0.84-1.28) | 0.99 (0.79-1.25) | 0.76 (0.57-1.02) |
| **Employment** |  |  |  |  |
| Unemployed | Reference | Reference | Reference | Reference |
| Paid employment | 0.79 (0.70-0.89) | 0.54 (0.48-0.62) | 0.35 (0.31-0.40) | 0.17 (0.13-0.21) |
| **Marital status** |  |  |  |  |
| Never married | Reference | Reference | Reference | Reference |
| Married/partner | 0.85 (0.69-1.04) | 0.90 (0.72-1.14) | 0.80 (0.62-1.03) | 0.82 (0.58-1.15) |
| Separated/divorced/widowed | 0.97 (0.77-1.23) | 1.14 (0.88-1.48) | 1.27 (0.96-1.68) | 1.41 (0.98-2.04) |

Abbreviation: MLTC, Multiple Long-Term Conditions

### Clusters of MLTC trajectory and all-cause mortality

The “Single-LTC” (aOR = 1.81; 95% CI 1.21 to 2.73), the “evolving MLTC” (aOR = 2.26; 95% CI 1.51 to 3.38), the “moderate MLTC” (aOR = 2.62; 95% CI 1.75 to 3.94), and the “high MLTC” (aOR = 4.03; 95% CI 2.64 to 6315) clusters were significantly associated with higher all-cause mortality, compared with the people in the “no-LTC” cluster (**Table 3**).

**Table 3.** Association between clusters of MLTC trajectory and all-cause mortality.

|  | Alive (14310, 95.6%) | Dead (652, 4.4%) | Unadjusted OR<br>(95%CI) | Adjusted<sup>1</sup> OR<br>(95%CI) | p-value<sup>2</sup> |
| --- | --- | --- | --- | --- | --- |
| **Trajectory cluster** |  |  |  |  |  |
| No-LTC | 2796 (98.9) | 30 (1.1) | Reference | Reference | <0.0001 |
| Single-LTC | 4668 (97.2) | 134 (2.8) | 2.69 (1.81-4.01) | 1.81 (1.21-2.73) |  |
| Evolving MLTC | 3566 (95.4) | 174 (4.6) | 4.59 (3.10-6.78) | 2.26 (1.51-3.38) |  |
| Moderate MLTC | 2349 (92.8) | 183 (7.2) | 7.22 (4.89-10.7) | 2.62 (1.75-3.94) |  |
| High MLTC | 931 (87.6) | 132 (12.4) | 13.6 (9.11-20.3) | 4.03 (2.64-6.15) |  |

<sup>1</sup>Adjusted for age, sex, ethnicity, education, employment status, and marital status. Age was included in the model as a squared term.
<sup>2</sup> p-value for trend.
Abbreviation: MLTC, multiple long-term condition.

## Discussion

This study examined clusters of MLTC based on the accumulation of conditions as trajectories over time, their associations with sociodemographic factors, and all-cause mortality among older adults in England. We identified five distinct clusters that can be described as “no-LTC”, “Single-LTC”, “evolving MLTC”, “moderate MLTC”, and “high MLTC”. We observed that the accumulation of MLTC over time progresses differently among older adults with distinction by sex, ethnicity, educational level, and employment status. Specifically, females and ethnic minorities showed faster/steeper progression towards increased numbers of MLTC, whereas higher education and paid employment had a protective effect on the increase of the accumulation of MLTC.

An interesting finding was that clusters with different initial levels and rates of change in MLTC indicating individual differences in the process of health deterioration. This is in line with previous studies that identified different rates of MLTC (25). Those with persistently high levels of multimorbidity have been also similarly identified in other population (26). However, consistent with the literature (25,26), we did not find any trajectories that indicated improvement in health over time (i.e., decreasing levels of MLTC). This may be due to the difficulty of recovery from long-term conditions among older adults.

The faster/steeper progression observed towards increased numbers of MLTC in females is in line with previous literature, which found that the accumulation of long-term conditions was more severe for older females (27). An explanation can be that females tend to live longer than males, and as a result, they are more likely to develop chronic conditions associated with ageing, such as arthritis and dementia. Clinicians should consider that females are at greater likelihood of MLTC. The faster development of MLTC in ethnic minorities can be explained by evidence suggesting that access and engagement with healthcare are limited for some population groups, often on the basis of ethnicity. Specifically, a review from NHS Race and Health Observatory (28) suggests that there are ‘clear barriers’ for people from minority ethnic backgrounds to seek help for mental health problems, and another research has also found lower access to cancer screening in the UK (29). Socioeconomic risk factors are known to be associated with MLTC (30). Our findings support the role of higher educational attainment, a major socioeconomic risk factor, on MLTC prevention. Targeting education inequality is expected to lead further to the restriction of worsening MLTC. The effect of educational attainment on MLTC is thought to be explained by other risk factors that may mediate this association, such as body mass index and smoking.

Over their life course, individuals develop MLTC. It is necessary to challenge the common statement that MLTC is inevitable in an ageing society. To do this, the focus on MLTC should shift from sole management of highrisk older individuals to include integrated population-level prevention strategies throughout the life course to address the drivers of MLTC. Programs that bridge multiple clinical specialities and healthcare units should be developed to focus on single individuals, their specific clinical profiles, and their specific clinical trajectories (31). Knowledge of how long-term conditions cluster, and especially how the status of MLTC can change over subsequent years, helps not only in understanding the complexity and dynamic evolution of MLTC clusters but also in supporting clinicians who manage co-occurring long-term conditions and health policymakers who plan care resources use.

This is the first study to examine trajectories of MLTC with a view to stratifying within MLTC to identify those at greatest risk among older adults in England. The main strength of the current study is the use of a large dataset, assessing longitudinal data to examine MLTC trajectories and a dataset that is nationally representative of people aged 50 years and older, including a wide range of long-term conditions and sociodemographics. However, the results of this study should be interpreted with some caution. First, the measurement of MLTC was limited to ten long-term conditions that was all of what was available in ELSA, which may not be exhaustive of all possible long-term conditions. Findings could be different if more long-term conditions are considered. Second, although we examined the correlates of MLTC trajectories using the variables measured at the baseline (wave 2), we cannot conclude on the directionality of the associations. Another limitation is that because our study utilised a longitudinal design that examined age-related changes, there may be inherent confounding of age and period effects. These effects could not be disentangled in this study due to the nature of our data. Lastly, the probability of being in a cluster membership is based on model assignment, which can lead to misclassification bias.

In conclusion, MLTC trajectories of older adults are characterised by dynamism but can still be tracked over time. Considering MLTC clusters will enable future researchers and practitioners to provide evidence in identifying older adults in England at a higher risk of worsening MLTC over time and further tailoring effective interventions for at-risk individuals. Targeting females and ethnic minorities is important for MLTC prevention. Higher levels of education can also lead to a further decrease in the number of long-term conditions. Policymakers should commit to increasing MLTC awareness.

## Supporting information

Strobe

## Data Availability

ELSA data were available through the UK Data Archive and are widely available to access in this way; as such our study data will not be made available for access.

## Acknowledgement

We would like to thank Ms Firoza Davies for her contribution as a patient and public representative in this study.

## References

1. World Health Organisation. Life tables [Internet]. 2022 [cited 2023 Mar 28]. Available from: https://www.who.int/data/gho/data/themes/mortality-and-global-health-estimates/ghe-life-expectancy-and-healthy-life-expectancy

2. Kingston A, Comas-Herrera A, Jagger C. Forecasting the care needs of the older population in England over the next 20 years: estimates from the Population Ageing and Care Simulation (PACSim) modelling study. Lancet Public Health. 2018 Sep;3(9):e447.

3. World Health Organisation. Ageing and health [Internet]. 2022 [cited 2023 Mar 28]. Available from: https://www.who.int/news-room/fact-sheets/detail/ageing-and-health

4. Ho ISS, Azcoaga-Lorenzo A, Akbari A, Davies J, Hodgins P, Khunti K, et al. Variation in the estimated prevalence of multimorbidity: systematic review and meta-analysis of 193 international studies. BMJ Open. 2022 Apr;12(4).

5. Makovski TT, Schmitz S, Zeegers MP, Stranges S, van den Akker M. Multimorbidity and quality of life: Systematic literature review and meta-analysis. Ageing Res Rev. 2019;53(April):100903.

6. Wang L, Si L, Cocker F, Palmer AJ, Sanderson K. A Systematic Review of Cost-of-Illness Studies of Multimorbidity. Appl Health Econ Health Policy. 2018;16(1):15–29.

7. Pathirana TI, Jackson CA. Socioeconomic status and multimorbidity: a systematic review and meta-analysis. Aust N Z J Public Health. 2018 Apr;42(2):186–94.

8. Nunes BP, Flores TR, Mielke GI, Thumé E, Facchini LA. Multimorbidity and mortality in older adults: A systematic review and meta-analysis. Arch Gerontol Geriatr. 2016 Nov;67:130–8.

9. Wang D, Li D, Mishra SR, Lim C, Dai X, Chen S, et al. Association between marital relationship and multimorbidity in middle-aged adults: a longitudinal study across the US, UK, Europe, and China. Maturitas. 2022 Jan;155:32–9.

10. Nguyen H, Wu YT, Dregan A, Vitoratou S, Chua KC, Prina AM. Multimorbidity patterns, all-cause mortality and healthy aging in older English adults: Results from the English Longitudinal Study of Aging. Geriatr Gerontol Int. 2020 Dec;20(12):1126–32.

11. Larsen FB, Pedersen MH, Friis K, Gluèmer C, Lasgaard M. A Latent Class Analysis of Multimorbidity and the Relationship to Socio-Demographic Factors and Health-Related Quality of Life. A National Population-Based Study of 162,283 Danish Adults. PLoS One. 2017 Jan;12(1):e0169426.

12. Fabbri E, Zoli M, Gonzalez-Freire M, Salive ME, Studenski SA, Ferrucci L. Aging and Multimorbidity: New Tasks, Priorities, and Frontiers for Integrated Gerontological and Clinical Research. J Am Med Dir Assoc. 2015 Aug;16(8):640.

13. Steptoe A, Breeze E, Banks J, Nazroo J. Cohort profile: the English longitudinal study of ageing. Int J Epidemiol. 2013 Dec;42(6):1640–8.

14. Banks J, Batty GD, Breedvelt JJF, Coughlin K, Crawford. R, Marmot M, et al. English Longitudinal Study of Ageing: Waves 0-9, 1998-2019 [data collection]. 3rd ed. UK Data Service. SN: 5050; 2021.

15. Cadar D, Abell J, Matthews FE, Brayne C, David Batty G, Llewellyn DJ, et al. Cohort Profile Update: The Harmonised Cognitive Assessment Protocol Sub-study of the English Longitudinal Study of Ageing (ELSA-HCAP). Int J Epidemiol. 2021 Jun;50(3):725–726I.

16. Dambha-Miller H, Simpson G, Hobson L, Olaniyan D, Hodgson S, Roderick P, et al. Integrating primary care and social services for older adults with multimorbidity: a qualitative study. Br J Gen Pract. 2021 Oct;71(711):E753–61.

17. Guthrie B, Payne K, Alderson P, McMurdo MET, Mercer SW. Adapting clinical guidelines to take account of multimorbidity. BMJ. 2012 Oct;345(7878).

18. Muggli E, Hearps S, Halliday J, Elliott EJ, Penington A, Thompson DK, et al. A data driven approach to identify trajectories of prenatal alcohol consumption in an Australian population-based cohort of pregnant women. Scientific Reports 2022 12:1. 2022 Mar;12(1):1–9.

19. Dalrymple K V., Vogel C, Godfrey KM, Baird J, Harvey NC, Hanson MA, et al. Longitudinal dietary trajectories from preconception to mid-childhood in women and children in the Southampton Women’s Survey and their relation to offspring adiposity: a group-based trajectory modelling approach. International Journal of Obesity 2021 46:4. 2021 Dec;46(4):758–66.

20. Mésidor M, Rousseau MC, O’Loughlin J, Sylvestre MP. Does group-based trajectory modeling estimate spurious trajectories? BMC Med Res Methodol. 2022 Dec;22(1).

21. Chen H, Zhou Y, Huang L, Xu X, Yuan C. Multimorbidity burden and developmental trajectory in relation to later-life dementia: A prospective study. Alzheimer’s & Dementia. 2022;1–10.

22. Quiñones AR, Nagel CL, Botoseneanu A, Newsom JT, Dorr DA, Kaye J, et al. Multidimensional trajectories of multimorbidity, functional status, cognitive performance, and depressive symptoms among diverse groups of older adults. Journal of Multimorbidity and Comorbidity. 2022 Nov;12:263355652211430.

23. Zhou H, Zhang H, Zhan Q, Bai Y, Liu S, Yang X, et al. Blood pressure trajectories in early adulthood and myocardial structure and function in later life. ESC Heart Fail. 2022 Apr;9(2):1258–68.

24. Walsh CA, Cahir C, Bennett KE. Longitudinal Medication Adherence in Older Adults With Multimorbidity and Association With Health Care Utilization: Results From the Irish Longitudinal Study on Ageing. Ann Pharmacother. 2021 Jan;55(1):5–14.

25. Strauss VY, Jones PW, Kadam UT, Jordan KP. Distinct trajectories of multimorbidity in primary care were identified using latent class growth analysis. J Clin Epidemiol. 2014;67(10):1163–71.

26. Lee SA, Joo S, Chai HW, Jun HJ. Patterns of multimorbidity trajectories and their correlates among Korean older adults. Age Ageing. 2021;50(4):1336–41.

27. García-Esquinas E, Ortolá R, Prina M, Stefler D, Rodríguez-Artalejo F, Pastor-Barriuso R. Trajectories of Accumulation of Health Deficits in Older Adults: Are There Variations According to Health Domains? J Am Med Dir Assoc. 2019;20(6):710–717.e6.

28. Kapadia D, Zhang J, Salway S, Nazroo J, Booth A, Villarroel-Williams N, et al. Ethnic Inequalities in Healthcare: A Rapid Evidence Review. 2022.

29. Szczepura A. Access to health care for ethnic minority populations. Vol. 81, Postgraduate Medical Journal. 2005. p. 141–7.

30. Ingram E, Ledden S, Beardon S, Gomes M, Hogarth S, Mcdonald H, et al. Household and area-level social determinants of multimorbidity: a systematic review. J Epidemiol Community Health (1978). 2021;75(3):232–41.

31. Hemingway H, Croft P, Perel P, Hayden JA, Abrams K, Timmis A, et al. Prognosis research strategy (PROGRESS) 1: A framework for researching clinical outcomes. BMJ (Online). 2013;346.

