## Supplementary tables for "Trajectories of multiple long-term conditions and mortality in older adults: A retrospective cohort study using English Longitudinal Study of Ageing (ELSA)"

### Supplements

**Supplementary Table 1**. Statistical parameters of the optimal number of clusters selection.

| **Number of groups** | **Group membership** | | **Trajectory shapes** | **BIC (sample size=15085)** | **APPA** | **OCC** |
| --- | --- | --- | --- | --- | --- | --- |
| **1** | (1) | 100 | 3 | -85493.21 | 1 | N/A |
| **2** | (1) | 53.49 | 33 | -73870.19 | 0.94 | 12.80 |
|  | (2) | 46.51 |  |  | 0.94 | 17.71 |
| **3** | (1) | 21.77 | 333 | -63524.35 | 0.97 | 105.25 |
|  | (2) | 53.83 |  |  | 0.96 | 18.03 |
|  | (3) | 24.40 |  |  | 0.95 | 67.92 |
| **4** | (1) | 19.24 | 3333 | -59262.14 | 0.96 | 93.69 |
|  | (2) | 36.07 |  |  | 0.93 | 24.44 |
|  | (3) | 32 |  |  | 0.90 | 19.03 |
|  | (4) | 12.69 |  |  | 0.96 | 172.34 |
| **5** | (1) | 19.35 | 33333 | -56474.28 | 0.97 | 119.07 |
|  | (2) | 30.77 |  |  | 0.90 | 18.95 |
|  | (3) | 25.43 |  |  | 0.88 | 23.76 |
|  | (4) | 17.15 |  |  | 0.90 | 44.02 |
|  | (5) | 7.31 |  |  | 0.95 | 284.50 |
| **5** | (1) | 18.57 | 03311 | -57000.83 | 0.96 | 109.69 |
|  | (2) | 31.21 |  |  | 0.90 | 19.32 |
|  | (3) | 25.82 |  |  | 0.87 | 20.91 |
|  | (4) | 17.12 |  |  | 0.90 | 44.32 |
|  | (5) | 7.27 |  |  | 0.95 | 259.29 |

*Note: Trajectory shapes (0=intercept, 1=linear, 2=quadratic, 3=cubic); BIC = Bayesian Information Criterion; APPA = average posterior probability assignment; OCC = odds of a correct classification according to maximum posterior probability group.*
